# Greater Absolute DTI-ALPS Asymmetry Is Associated with Higher Risk of Conversion to Alzheimer Disease

**DOI:** 10.64898/2026.11.27.26369020

**Authors:** Jun Hyeok Choi, Jun-Hee Kim, Hyochul Lee, Seung Hong Choi, Roh-Eul Yoo, the Alzheimer’s Disease Neuroimaging Initiative

## Abstract

**Background:** Diffusion tensor imaging (DTI) analysis along the perivascular space (ALPS) is conventionally summarized by averaging left- and right-hemisphere measurements, which may obscure interhemispheric diffusion heterogeneity relevant to Alzheimer disease (AD) conversion.

**Purpose:** To determine whether absolute ALPS asymmetry is associated with subsequent AD conversion beyond bilateral mean ALPS and structural MRI markers.

**Materials and Methods:** This retrospective Alzheimer’s Disease Neuroimaging Initiative study included cognitively normal participants and participants with mild cognitive impairment who underwent 3-T diffusion MRI between June 2010 and September 2025; follow-up continued through July 2026. Absolute ALPS asymmetry was the absolute left-right difference in the ALPS index. Diagnosis-stratified Cox models adjusted for age, sex, education, baseline Mini-Mental State Examination score, log-transformed white matter hyperintensity volume, brain parenchymal fraction, and bilateral mean ALPS. A likelihood-ratio test assessed incremental model fit. Sensitivity analyses evaluated alternative asymmetry definitions, structural and diffusion correlates, and technical covariates.

**Results:** Among 1,131 participants (mean age, 72 years ± 8 [SD]; 607 women), 115 converted to AD. In an unadjusted comparison, converters had higher baseline absolute ALPS asymmetry than nonconverters (mean difference, 0.021; 95% confidence interval [CI]: 0.001, 0.041; P = .04). Each 1 SD increase in absolute ALPS asymmetry was associated with conversion (adjusted hazard ratio [HR], 1.26; 95% CI: 1.07, 1.48; P = .005), and its addition improved model fit (likelihood-ratio χ² = 6.43; P = .01). The adjusted 60-month risk difference between the cohort mean and 1 SD above the mean was 2.3 percentage points (95% CI: 0.54, 4.02). Bilateral mean ALPS was not associated with conversion in the same model (HR, 0.94; 95% CI: 0.76, 1.16; P = .55); hemisphere-specific ALPS and the signed ALPS difference also showed no association.

**Conclusion:** Absolute ALPS asymmetry provided prognostic information beyond bilateral mean ALPS and structural MRI markers for subsequent AD conversion.

**Summary Statement:** Absolute asymmetry in the diffusion MRI-derived analysis along the perivascular space (ALPS) index was associated with Alzheimer disease conversion after adjustment for clinical measures, structural MRI, and bilateral mean ALPS.

**Key Results:**

- Among 1,131 participants (115 Alzheimer disease conversions), each standard deviation increase in absolute ALPS asymmetry was associated with conversion (adjusted hazard ratio, 1.26; P = .005) and improved model fit (P = .01).
- Bilateral mean ALPS, hemisphere-specific ALPS, and the signed ALPS difference showed no association with Alzheimer disease conversion.
- Compared with absolute ALPS asymmetry set to the cohort mean, setting it one standard deviation higher corresponded to 1.6- and 2.3-percentage-point increases in adjusted Alzheimer disease conversion risk at 36 and 60 months, respectively.

## Introduction

The glymphatic system has been proposed to clear metabolic waste through cerebrospinal-interstitial fluid exchange (1–4). Impaired clearance of amyloid-β and tau may contribute to neurodegeneration and has been linked to Alzheimer disease (AD) and other neurodegenerative diseases (4–10).

Direct assessment of glymphatic clearance in humans generally requires intrathecal contrast administration; therefore, noninvasive MRI markers have been developed. Diffusion tensor imaging (DTI) analysis along the perivascular space (ALPS), or DTI-ALPS, combines directional diffusivities in periventricular white matter, where perivascular spaces run perpendicular to major fiber tracts (5,11). Previous longitudinal studies have linked lower ALPS levels to subsequent clinical progression and conversion from mild cognitive impairment (MCI) to AD (9,10,12). A noninvasive marker that complements clinical information could support risk stratification and follow-up planning (13).

AD pathology can develop asymmetrically across hemispheres. Amyloid-β laterality has been associated with subsequent changes in tau laterality, suggesting hemispheric differences in vulnerability to pathological accumulation (14). Perivascular transport also shows regional organization and activity-dependent regulation, providing a biological basis for investigating interhemispheric differences (15,16). Greater ALPS asymmetry has been associated with unfavorable seizure outcomes in MRI-negative focal epilepsy, supporting the potential prognostic relevance of bilateral differences in another neurological disorder (17). Together, these observations motivate investigation of ALPS asymmetry as a candidate marker of regional vulnerability in AD. Most DTI-ALPS studies summarize bilateral measurements by averaging, which may obscure such differences. Although previous cross-sectional work evaluated signed right-left ALPS differences across cognitively normal (CN), MCI, and AD groups, no significant between-group differences were observed (18). Whether the magnitude of interhemispheric ALPS asymmetry provides prognostic information for AD conversion beyond bilateral mean ALPS and established clinical and structural MRI measures remains unclear.

We hypothesized that absolute interhemispheric DTI-ALPS asymmetry (absolute ALPS asymmetry) would provide prognostic information beyond clinical and structural MRI measures. Hemisphere-specific ALPS and the signed ALPS difference (left minus right) were examined to distinguish asymmetry magnitude from direction. Additional analyses evaluated alternative definitions, structural and diffusion correlates, and prediction performance. We evaluated whether absolute ALPS asymmetry was associated with AD conversion after adjustment for clinical variables, white matter hyperintensity (WMH) volume, brain parenchymal fraction (BPF), and bilateral mean ALPS in the longitudinal Alzheimer’s Disease Neuroimaging Initiative (ADNI) cohort (19).

## Materials and Methods

### Study Design and Participants

Data used in the preparation of this article were obtained from the Alzheimer’s Disease Neuroimaging Initiative (ADNI) database (adni.loni.usc.edu). The ADNI was launched in 2003 as a public-private partnership, led by Principal Investigator Michael W. Weiner, MD. The primary goal of ADNI has been to test whether serial magnetic resonance imaging (MRI), positron emission tomography (PET), other biological markers, and clinical and neuropsychological assessment can be combined to measure the progression of mild cognitive impairment (MCI) and early Alzheimer’s disease (AD).

This retrospective longitudinal study used deidentified ADNI data from ADNI-1, ADNI-GO, ADNI-2, ADNI-3, and ADNI-4. Each participating site obtained institutional review board approval, and participants or authorized representatives provided written informed consent. Reporting followed the Strengthening the Reporting of Observational Studies in Epidemiology (STROBE) recommendations.

Eligible participants were CN or had MCI at baseline and had subsequent diagnostic follow-up, an available Mini-Mental State Examination (MMSE) score, and structural MRI data. Participants were excluded for the absence of a diagnosis within the baseline window, AD at baseline, no post-index follow-up, unavailable MMSE or structural MRI data, fewer than 30 diffusion directions, or ALPS processing failure (Figure 1). Study baseline was each participant’s first usable diffusion MRI scan. Baseline diagnosis was the assessment closest to the scan within ±90 days. Incident AD was the first subsequent site-adjudicated AD diagnosis. Follow-up extended from baseline MRI to AD diagnosis or the last diagnostic assessment. All eligible participants were included without an a priori sample-size calculation. Race and ethnicity were unavailable. Details for the data source, baseline diffusion MRI, clinical variables and outcome are provided in the Supplemental Materials and Methods.

**Figure 1.**
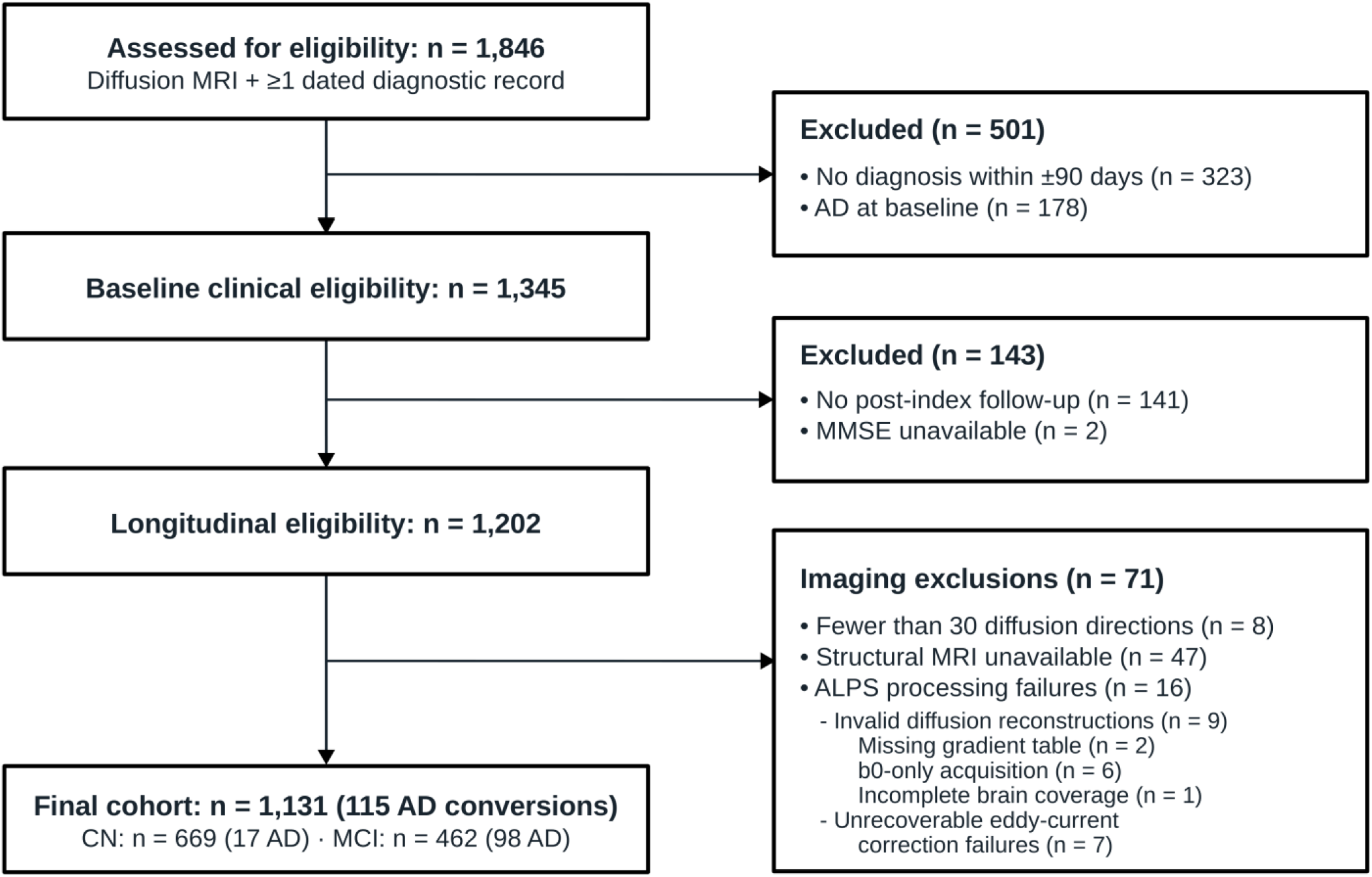
Flowchart of participant inclusion and exclusion. Sequential clinical, longitudinal, and imaging eligibility criteria yielded 1,131 participants. AD conversion occurred in 115 participants, including 17 of 669 with CN and 98 of 462 with MCI. The 16 ALPS processing failures comprised nine invalid diffusion reconstructions and seven unrecoverable eddy-current correction failures. AD = Alzheimer disease; ALPS = analysis along the perivascular space; CN = cognitively normal; MCI = mild cognitive impairment; MMSE = Mini-Mental State Examination.

### MRI Acquisition, Processing, and ALPS Measures

BPF and WMH volume were obtained from ADNI-provided University of California, Davis four-tissue segmentation of T1-weighted and coregistered fluid-attenuated inversion recovery (FLAIR) images. Left and right ventricular, cortical gray matter, and cortical white matter volumes for structural asymmetry analyses came from the ADNI UCSFFSX7 FreeSurfer 7.x dataset.

Diffusion MRI was acquired on 3-T Siemens, GE Healthcare, and Philips systems. Single-shell and multi-shell acquisitions were defined by the number of diffusion shells at b ≥ 50 s/mm². Automated DTI-ALPS processing followed a standardized pipeline using FMRIB Software Library (FSL) 6.0 and MRtrix3 3.0.8 (20,21), including denoising, Gibbs-ringing correction, eddy-current and motion correction, diffusion-tensor fitting, and affine transformation of tensor maps to the JHU-ICBM-FA 1-mm template (FA = fractional anisotropy) using FMRIB’s Linear Image Registration Tool (FLIRT) and applywarp in FSL (20) (Figure 2A, Supplemental Materials and Methods).

**Figure 2.**
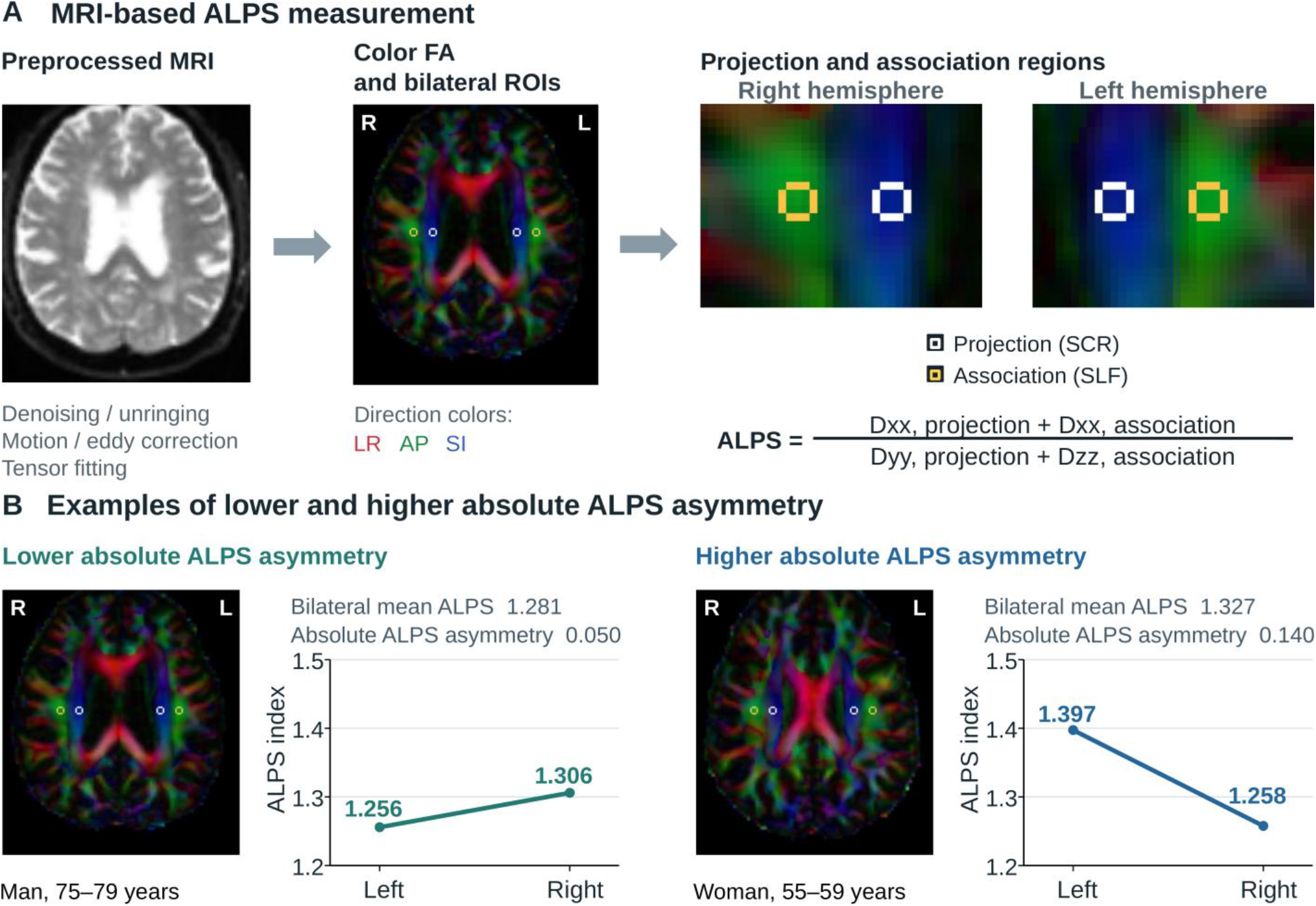
MRI-based ALPS measurement and examples of absolute ALPS asymmetry. A, Diffusion MRI preprocessing, axial color-coded FA images, and the bilateral projection and association ROIs used to calculate ALPS. Dxx, Dyy, and Dzz are diagonal diffusion-tensor components along the left-right (LR), anterior-posterior (AP), and superior-inferior (SI) axes. **B,** Axial color-coded FA images and bilateral ROI overlays for two participants with lower and higher absolute ALPS asymmetry. The lower-asymmetry example (man aged 75–79 years) had a bilateral mean ALPS of 1.281 and an absolute ALPS asymmetry of 0.050. The higher-asymmetry example (woman aged 55– 59 years) had corresponding values of 1.327 and 0.140. Means and absolute differences were calculated from unrounded values. ALPS = analysis along the perivascular space; FA = fractional anisotropy; L = left; R = right; ROI = region of interest; SCR = superior corona radiata; SLF = superior longitudinal fasciculus.

Four spherical regions of interest (ROIs) (2.5-mm radius) sampled bilateral projection and association white matter adjacent to the lateral ventricular bodies. Hemispheric ALPS was the ratio of summed x-axis diffusivities in projection and association fibers to summed y-axis projection-fiber and z-axis association-fiber diffusivities (Eq. 1).

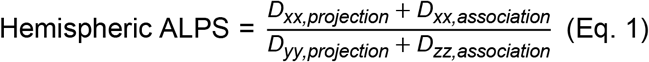

Bilateral mean ALPS was the average of hemispheric values; signed ALPS difference was left minus right. The primary exposure, absolute ALPS asymmetry, was the absolute left-right difference (|ALPS_L_ − ALPS_R_|) (Eq. 2; Figure 2B).

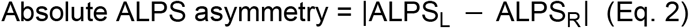

Two alternative asymmetry definitions were evaluated: the absolute left-right difference divided by bilateral mean ALPS (mean-normalized relative asymmetry) and the absolute log ratio of left to right ALPS.

Outliers more than 3 standard deviations (SDs) from the mean in regional diffusivities or ALPS indices were visually reviewed for ROI placement, symmetry, and image integrity. Details for the ALPS calculation and quality control are provided in the Supplemental Materials and Methods.

### Structural MRI and Technical Variables

Structural covariates were BPF and WMH volume. BPF was calculated as the sum of gray matter, normal-appearing white matter, and WMH volumes divided by intracranial volume. Normal-appearing white matter referred to the white matter compartment excluding voxels classified as WMH in the four-tissue segmentation. WMH volume was modeled as ln(1 + WMH volume) (Supplemental Materials and Methods).

Technical sensitivity models adjusted for scanner manufacturer, diffusion directions, section thickness, repetition time, and echo time. A comprehensive model additionally included within-scan motion, registration-derived head orientation, and acquisition-plane orientation derived from Digital Imaging and Communications in Medicine (DICOM) data.

### Statistical Analysis

Analyses used Python 3.12.4, lifelines 0.30.0, and pycox 0.3.0. Continuous variables were summarized as mean ± SD or median (interquartile range), and categorical variables as counts and percentages. Baseline characteristics were compared by observed AD conversion status using two-sided Welch t tests for means, Mann-Whitney U tests for WMH volume, and Pearson χ² tests for sex and baseline diagnosis. Baseline CN and MCI groups were compared using the same tests for the corresponding variables.

An unadjusted two-sided Welch t test compared baseline absolute ALPS asymmetry between participants who converted to AD during observed follow-up and those without documented AD conversion through their last diagnostic assessment. The group comparisons were descriptive and did not account for follow-up duration or censoring.

The reference Cox model (22) included age, sex, education, baseline MMSE, log-transformed WMH volume, BPF, and bilateral mean ALPS. The primary Cox model added absolute ALPS asymmetry. Both models were stratified by baseline diagnosis and used the same participants, with follow-up in days and continuous predictors standardized by the cohort SD. Robust standard errors were used for hazard ratio (HR) confidence intervals (CIs) and two-sided Wald tests. A likelihood-ratio (LR) test with 1 degree of freedom (df) compared the primary Cox model with the reference model. LR tests for the association-fiber Dzz asymmetry analysis compared each expanded model with the specified reduced model on the same participants. We compared model fit using partial Akaike information criterion (AIC) and estimated the apparent Harrell C-index by pooling comparable pairs within diagnosis strata. Percentile 95% CIs for the C-index used 1,000 participant-level bootstrap samples with the fitted risk scores held fixed. Schoenfeld residuals assessed proportional hazards, and a three-knot restricted cubic spline assessed nonlinearity.

Adjusted conversion probabilities were estimated by marginal standardization, setting bilateral mean ALPS or absolute ALPS asymmetry to the cohort mean or 1 SD above it while retaining other observed covariates and diagnosis strata. Risk differences at 36 and 60 months used percentile 95% CIs from 500 participant-level bootstrap samples (Supplemental Materials and Methods).

Prediction analyses focused on conventional Cox regression and DeepSurv (23). Additional CoxTime and DeepHitSingle analyses (24,25) are reported in the Supplemental Materials and Methods. Reference covariates were age, sex, education, baseline MMSE, log-transformed WMH volume, BPF, and bilateral mean ALPS. Baseline diagnosis defined strata for Cox regression and was a predictor for DeepSurv. Extended models added absolute ALPS asymmetry. Both models used the same outer splits in five repetitions of five-fold cross-validation. Preprocessing was fitted within training data. We assessed time-dependent area under the receiver operating characteristic curve (AUC) and Brier scores at 36 and 60 months (26,27). Participant-level bootstrap resampling used fixed out-of-fold predictions and censoring weights. Cox AUC contrasts used 10,000 resamples and nominal P values. DeepSurv retained the multiplicity adjustments from the original three-model neural analysis.

Sensitivity analyses evaluated hemisphere-specific ALPS, the signed ALPS difference, the two alternative asymmetry definitions, and absolute asymmetries of the constituent tensor components. We also examined influential observations, structural asymmetry, and technical covariates, as described in the Supplemental Materials and Methods. Sensitivity analyses used nominal two-sided P values.

## Results

### Participant Characteristics

Among 1,846 participants assessed for eligibility, 323 were excluded because no diagnosis was available within ±90 days of baseline MRI, and 178 had AD at baseline, leaving 1,345 participants. A further 141 participants had no post-index follow-up, and two had unavailable MMSE scores, leaving 1,202 participants. Imaging exclusions comprised fewer than 30 diffusion directions (n = 8), unavailable structural MRI data (n = 47), and ALPS processing failures (n = 16). Processing failures included invalid diffusion reconstructions (n = 9) and unrecoverable eddy-current correction failures (n = 7). The final cohort included 1,131 participants (Figure 1; Table S1).

Mean age was 72.1 years ± 7.7 [SD], and 607 participants (53.7%) were women. The cohort included 669 participants with CN and 462 with MCI (Figure 1; Table S2).

Baseline diffusion MRI was acquired on 3-T systems from Siemens (n = 600), GE Healthcare (n = 379), or Philips (n = 152). Scanner model names were available for all 1,131 participants, and model-specific counts are provided in Table S3. There were 857 single-shell and 274 multi-shell acquisitions (diffusion-shell threshold, b ≥ 50 s/mm²). Acquisition parameters were heterogeneous (in-plane pixel spacing, 0.91–3.0 mm; section thickness, 2.0–4.0 mm; repetition time, 3,067–17,523 ms; echo time, 47.7–105.0 ms) (Supplemental Materials and Methods, Section 4).

Baseline scans were acquired from June 17, 2010, to September 30, 2025, and follow-up extended through July 14, 2026. During follow-up, 115 participants (10.2%) converted to AD, including 17 of 669 participants with CN (2.5%) and 98 of 462 participants with MCI (21.2%). Median follow-up was 36.3 months (interquartile range, 12.2–82.3 months) overall, 48.4 months (interquartile range, 12.5–88.1 months) in participants with CN, and 25.4 months (interquartile range, 11.6–58.7 months) in participants with MCI. Compared with nonconverters, converters were older, more often had baseline MCI, had lower baseline MMSE scores and BPF, and had greater WMH volume (Table 1).

**Table 1.** Baseline Participant Characteristics by AD Conversion Status.

| **Characteristic** | **Overall (n = 1,131)** | **Converters (n = 115)** | **Nonconverters (n = 1,016)** | **P** |
|---|---|---|---|---|
| Age, years | 72.1 ± 7.7 | 75.6 ± 6.7 | 71.7 ± 7.8 | < .001 |
| Women, n (%) | 607 (53.7) | 49 (42.6) | 558 (54.9) | .01 |
| Education, years | 16.4 ± 2.5 | 16.5 ± 2.6 | 16.4 ± 2.5 | .59 |
| Baseline MMSE | 28.4 ± 1.7 | 27.0 ± 2.2 | 28.6 ± 1.6 | < .001 |
| WMH volume, mL | 1.95 (0.61–5.45) | 4.76 (2.23–11.05) | 1.72 (0.54–4.88) | < .001 |
| BPF, % | 75.3 ± 2.9 | 73.7 ± 2.9 | 75.5 ± 2.8 | < .001 |
| Bilateral mean ALPS | 1.26 ± 0.19 | 1.20 ± 0.18 | 1.27 ± 0.19 | < .001 |
| Absolute ALPS asymmetry | 0.11 ± 0.09 | 0.13 ± 0.10 | 0.11 ± 0.09 | .04 |
| Baseline MCI, n (%) | 462 (40.8) | 98 (85.2) | 364 (35.8) | < .001 |

Continuous data are mean ± SD or median (IQR). P values compare observed AD converters with nonconverters using two-sided Welch t tests for means, the Mann-Whitney U test for WMH volume, and Pearson χ² tests for sex and baseline diagnosis. Nonconverters had no documented AD conversion through their last diagnostic assessment. AD = Alzheimer disease; ALPS = analysis along the perivascular space; BPF = brain parenchymal fraction; IQR = interquartile range; MCI = mild cognitive impairment; MMSE = Mini-Mental State Examination; SD = standard deviation; WMH = white matter hyperintensity.

### Difference in Absolute ALPS Asymmetry between AD Converters and Nonconverters

In an unadjusted comparison, participants who converted to AD during their available follow-up (n = 115) had higher baseline absolute ALPS asymmetry than nonconverters (n = 1,016) (mean difference, 0.021; 95% CI: 0.001, 0.041; P = .04) (Figure 3).

**Figure 3.**
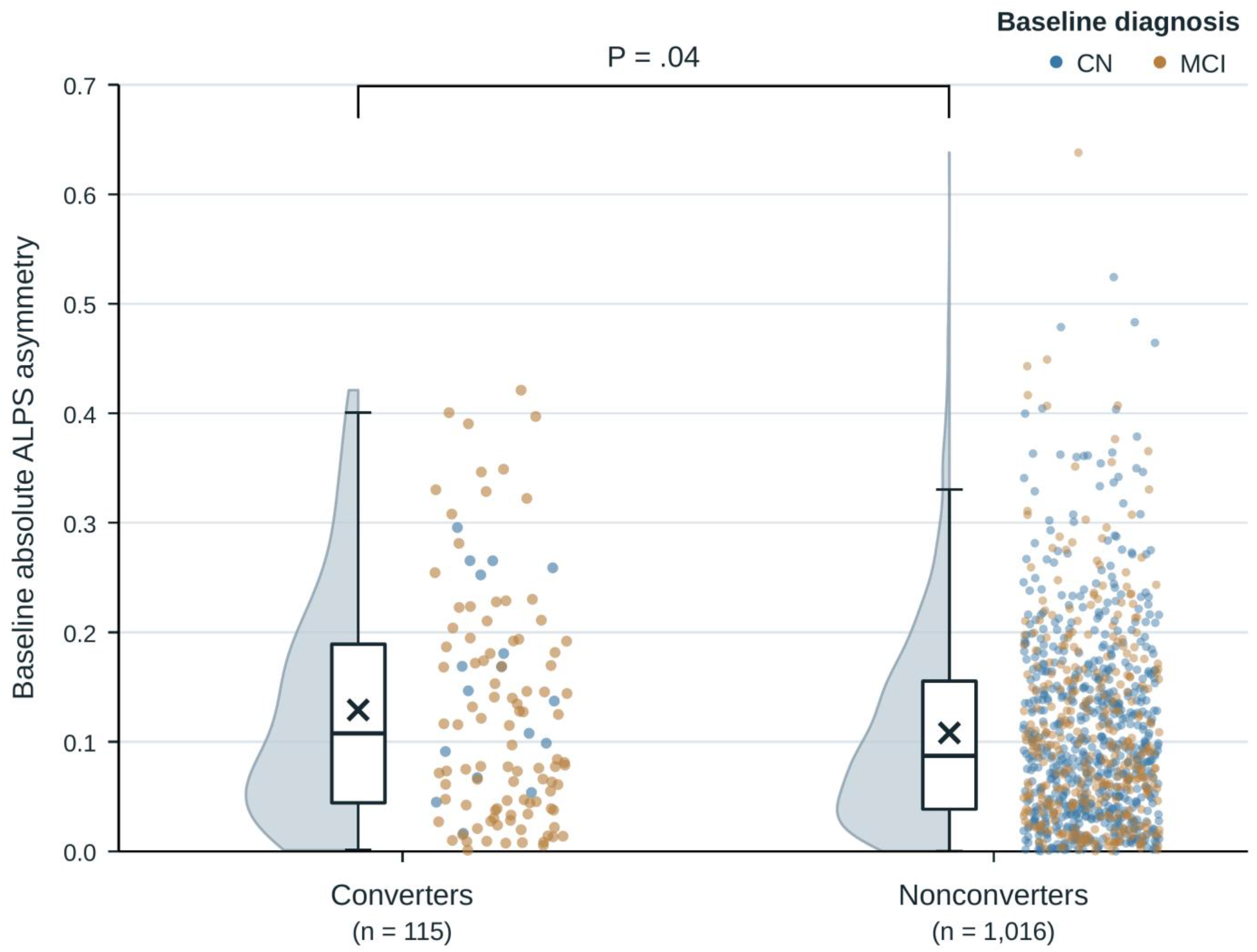
Baseline absolute ALPS asymmetry by observed AD conversion status. Baseline absolute ALPS asymmetry in converters (n = 115) and nonconverters (n = 1,016). Nonconverters had no documented AD conversion through their last diagnostic assessment. Half violins show the estimated distribution density. Boxes show the interquartile range and median, whiskers extend to the most extreme values within 1.5 times that range, and X symbols mark group means. Points show participants, colored by baseline diagnosis. The bracket marks the two-sided Welch t test, which did not account for follow-up duration or censoring. AD = Alzheimer disease; ALPS = analysis along the perivascular space; CI = confidence interval; CN = cognitively normal; MCI = mild cognitive impairment.

### Association between Absolute ALPS Asymmetry and AD Conversion Risk

Greater absolute ALPS asymmetry was associated with AD conversion (adjusted HR per 1 SD, 1.26; 95% CI: 1.07, 1.48; P = .005) (Table 2; Figure 4A). Adding absolute ALPS asymmetry to the clinical, structural MRI, and bilateral mean ALPS model improved fit (likelihood-ratio χ² = 6.43; P = .01) (Table 3). Apparent Harrell C-index was 0.745 (95% CI: 0.698, 0.789) for the reference model and 0.760 (95% CI: 0.716, 0.798) for the primary Cox model. Partial AIC was 1,121.03 and 1,116.60, respectively (Table 4). Bilateral mean ALPS was not associated with conversion when modeled with absolute ALPS asymmetry (HR, 0.94; 95% CI: 0.76, 1.16; P = .55) (Table 2; Figure 4A). In the primary Cox model, BPF (HR, 0.59; 95% CI: 0.45, 0.77; P < .001) and log-transformed WMH volume (HR, 1.37; 95% CI: 1.12, 1.68; P = .002) were associated with AD conversion (Table 2; Figure 4A). Baseline MMSE was also associated with AD conversion (HR, 0.59; 95% CI: 0.52, 0.68; P < .001) (Table 2).

**Figure 4.**
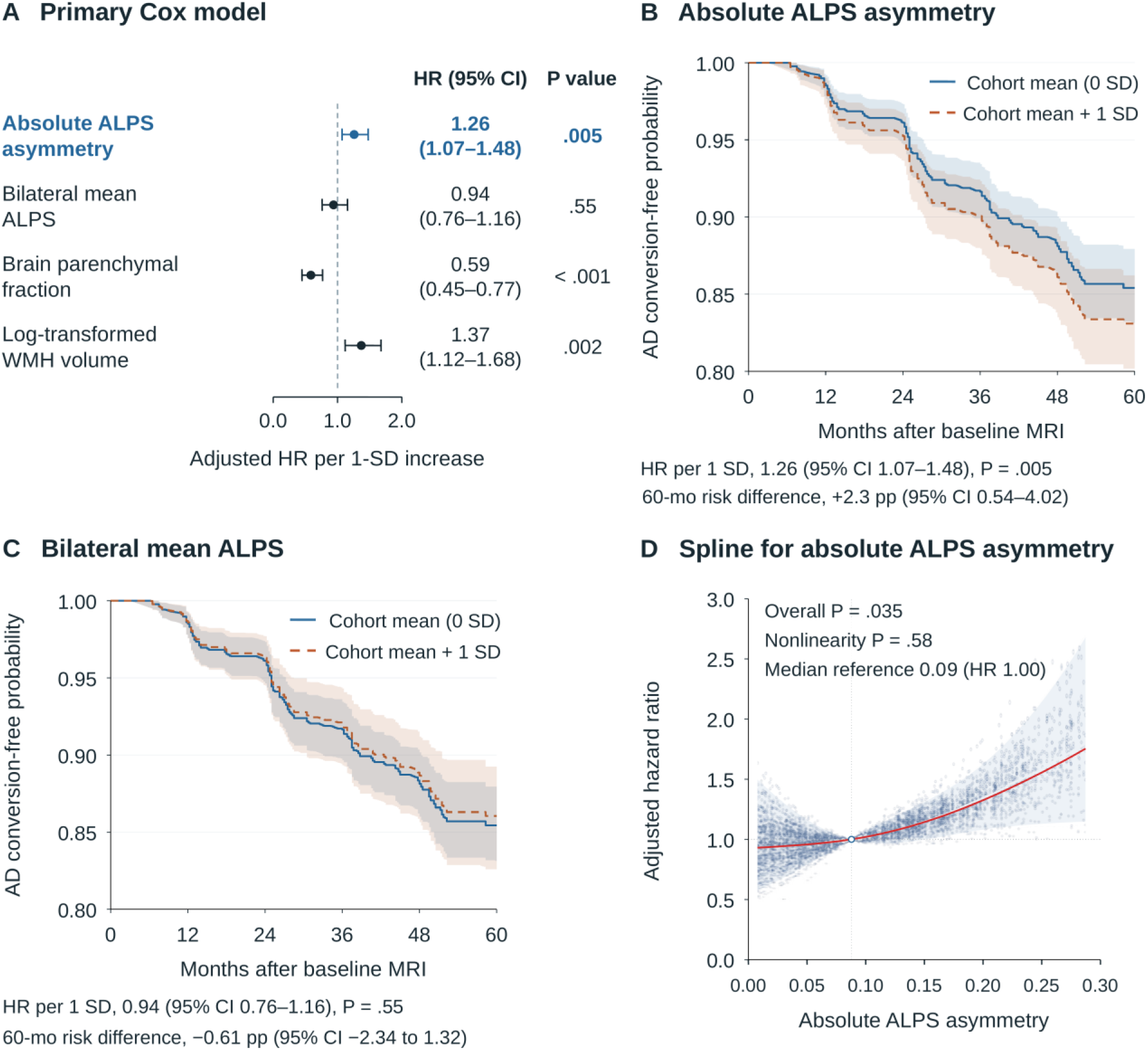
Primary Cox model estimates, adjusted probabilities, and spline association. A, Imaging covariates from the primary Cox model (mutually adjusted). Circles and horizontal bars show adjusted HRs per 1 SD increase and 95% CIs. P values are from two-sided Wald tests with robust standard errors. **B,** Absolute ALPS asymmetry. **C,** Bilateral mean ALPS. Marginally standardized AD conversion-free probability curves from the primary Cox model (n = 1,131) compare the cohort mean (0 SD, solid blue) with 1 SD above it (dashed orange), retaining other covariates and diagnosis strata at observed values. Bands show pointwise 95% bootstrap CIs from 500 resamples. Risk differences use unrounded conversion probabilities. **D,** Three-knot restricted cubic spline for absolute ALPS asymmetry over its 5th-95th percentiles. The fitted HR equals 1 at the median reference (0.088), marked by an open circle and vertical dotted line. Shading shows pointwise 95% CIs. AD = Alzheimer disease; ALPS = analysis along the perivascular space; CI = confidence interval; HR = hazard ratio; pp = percentage points; SD = standard deviation; WMH = white matter hyperintensity.

**Table 2.** Multivariable Cox Associations with AD Conversion.

| **Variable** | **Adjusted HR (95% CI)** | **P** |
|---|---|---|
| Absolute ALPS asymmetry | 1.26 (1.07, 1.48) | .005 |
| Bilateral mean ALPS | 0.94 (0.76, 1.16) | .55 |
| Age | 0.89 (0.66, 1.18) | .41 |
| Female sex (vs male) | 1.17 (0.80, 1.70) | .42 |
| Education | 1.14 (0.94, 1.38) | .17 |
| Baseline MMSE | 0.59 (0.52, 0.68) | < .001 |
| Log-transformed WMH volume | 1.37 (1.12, 1.68) | .002 |
| BPF | 0.59 (0.45, 0.77) | < .001 |

The primary Cox model included age, sex, education, baseline MMSE, log-transformed WMH volume, BPF, bilateral mean ALPS, and absolute ALPS asymmetry and was stratified by baseline diagnosis. All eight fitted covariate coefficients are shown (n = 1,131; 115 events). Continuous-variable HRs are per 1 SD; the sex HR compares female with male participants. HRs use robust standard errors. Baseline diagnosis defines separate baseline hazards and has no single fitted HR. WMH volume was modeled as ln(1 + WMH volume). AD = Alzheimer disease; ALPS = analysis along the perivascular space; BPF = brain parenchymal fraction; CI = confidence interval; HR = hazard ratio; MMSE = Mini-Mental State Examination; SD = standard deviation; WMH = white matter hyperintensity.

**Table 3.** Nested Likelihood-Ratio Tests of Cox Models.

| **Reduced model** | **Added Variable(s)** | **df** | **LR χ²** | **P** |
|---|---|---|---|---|
| **Single-marker addition** |  |  |  |  |
| Reference model | + Absolute ALPS asymmetry | 1 | 6.43 | .01 |
| Reference model | + Association-fiber Dzz asymmetry | 1 | 6.46 | .01 |
| **Joint and incremental addition** |  |  |  |  |
| Reference model | + Both asymmetry measures | 2 | 9.60 | .008 |
| Reference + Association-fiber Dzz asymmetry | + Absolute ALPS asymmetry | 1 | 3.15 | .08 |
| Reference + Absolute ALPS asymmetry | + Association-fiber Dzz asymmetry | 1 | 3.17 | .07 |

The reference Cox model included age, sex, education, baseline MMSE, log-transformed WMH, BPF, and bilateral mean ALPS. All models were stratified by baseline diagnosis and fitted in the same 1,131 participants (115 AD conversions). Absolute ALPS asymmetry and association-fiber Dzz asymmetry were defined as absolute left-right differences and standardized using full-cohort means and standard deviations. One-degree-of-freedom likelihood-ratio tests evaluated the addition of one asymmetry measure relative to the specified reduced model. The 2-df test compared the joint model with the reference model. AD = Alzheimer disease; ALPS = analysis along the perivascular space; BPF = brain parenchymal fraction; df = degrees of freedom; Dzz = z-axis diffusivity measured in the association-fiber region of interest; LR = likelihood-ratio; MMSE = Mini-Mental State Examination; WMH = white matter hyperintensity.

**Table 4.** Harrell C-index and Partial AIC for Cox Models.

| **Model** | **Apparent C-index (95% CI)** | **Partial AIC** |
|---|---|---|
| Reference model | 0.745 (0.698, 0.789) | 1,121.03 |
| Reference + absolute ALPS asymmetry | 0.760 (0.716, 0.798) | 1,116.60 |
| Reference + association-fiber Dzz asymmetry | 0.750 (0.702, 0.793) | 1,116.57 |
| Reference + both asymmetry measures | 0.762 (0.717, 0.800) | 1,115.42 |

The reference Cox model included age, sex, education, baseline MMSE, log-transformed WMH volume, BPF, and bilateral mean ALPS. All models were stratified by baseline diagnosis and fitted in the same 1,131 participants (115 AD conversions). Absolute ALPS asymmetry and association-fiber Dzz asymmetry were defined as absolute left-right differences and standardized using full-cohort means and standard deviations. Apparent Harrell C-index was calculated from comparable pairs within diagnosis strata. Percentile 95% CIs used 1,000 participant-level bootstrap resamples with fitted risk scores held fixed. Akaike information criterion was calculated from each Cox model’s partial log-likelihood. AD = Alzheimer disease; AIC = Akaike information criterion; ALPS = analysis along the perivascular space; BPF = brain parenchymal fraction; CI = confidence interval; Dzz = z-axis diffusivity measured in the association-fiber region of interest; MMSE = Mini-Mental State Examination; WMH = white matter hyperintensity.

At 36 months, standardized conversion probabilities were 8.4% (95% CI: 6.57%, 10.19%) when absolute ALPS asymmetry was set to the cohort mean and 9.9% (95% CI: 7.68%, 12.06%) when it was set to 1 SD above the mean, a 1.6-percentage-point difference (95% CI: 0.34, 2.72) (Figure 4B). Corresponding 60-month probabilities were 14.6% (95% CI: 12.06%, 16.96%) and 16.9% (95% CI: 13.79%, 19.84%), a 2.3-percentage-point difference (95% CI: 0.54, 4.02) (Figure 4B). Risk differences were calculated from unrounded probabilities. Bilateral mean ALPS had a 60-month difference of −0.61 percentage points (95% CI: −2.34, 1.32) (Figure 4C). The spline showed an overall association with AD conversion (P = .035), with no evidence of nonlinearity (P = .58; Figure 4D), and Schoenfeld residuals supported proportional hazards.

### AD Conversion Prediction Performance

For conventional Cox regression, reference and extended AUCs were 0.9084 and 0.9093 at 36 months (difference, +0.09 percentage points; 95% CI: −0.57, 0.77; P = .78) and 0.9141 and 0.9178 at 60 months (difference, +0.38 percentage points; 95% CI: −0.32, 1.13; P = .31). For DeepSurv, AUCs were 0.9055 and 0.9059 at 36 months (difference, +0.05 percentage points; 95% CI: −0.72, 0.83; adjusted P = .93) and 0.9014 and 0.9052 at 60 months (difference, +0.38 percentage points; 95% CI: −0.55, 1.54; adjusted P > .99) (Table S4). DeepSurv Brier-score contrasts are reported in Table S5.

### Sensitivity Analyses

Left ALPS (HR, 0.98; 95% CI: 0.80, 1.21; P = .87), right ALPS (HR, 0.97; 95% CI: 0.79, 1.20; P = .79), and the signed ALPS difference (HR, 1.02; 95% CI: 0.82, 1.25; P = .89) were not associated with AD conversion (Figure 5A, Table S6). Bilateral mean ALPS was not associated with conversion when modeled without absolute ALPS asymmetry (HR, 0.98; 95% CI: 0.80, 1.20; P = .81) (Table S6). Mean-normalized relative asymmetry and the absolute log ratio yielded similar HRs (1.22; Wald P = .01 for both) (Table S6).

**Figure 5.**
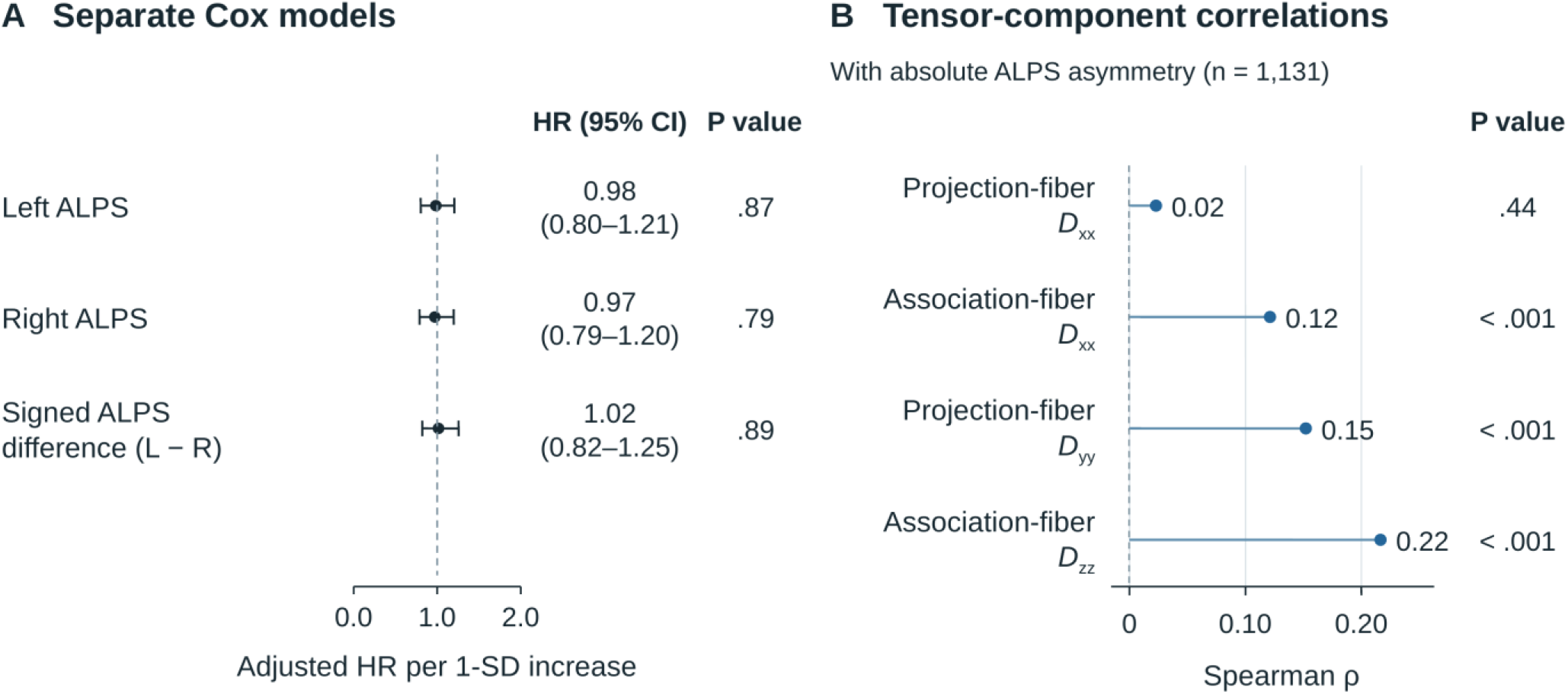
Hemisphere-specific ALPS measures and tensor component asymmetry. A, Separate Cox models for left ALPS, right ALPS, and the signed ALPS difference (left minus right). Only the signed-difference model additionally adjusted for bilateral mean ALPS. Circles and horizontal bars show adjusted HRs per 1 SD increase and 95% CIs. P values are from two-sided Wald tests with robust standard errors. **B,** Spearman correlations between absolute ALPS asymmetry and each tensor component’s absolute left-right difference (n = 1,131). P values are two-sided and unadjusted. AD = Alzheimer disease; ALPS = analysis along the perivascular space; CI = confidence interval; HR = hazard ratio; SD = standard deviation.

Association-fiber Dzz asymmetry had the strongest correlation with absolute ALPS asymmetry among the four tensor components (Spearman ρ = 0.22; P < .001) (Figure 5B). With both measures in the model, the HR for absolute ALPS asymmetry was 1.19 (95% CI: 0.99, 1.43; P = .07) (Table S7). Their joint addition improved fit (2-df LR χ² = 9.60; P = .008), and the two separate 1-df tests for incremental contributions yielded P = .08 for adding absolute ALPS asymmetry and P = .07 for adding association-fiber Dzz asymmetry (Table 3).

Additional structural MRI, technical covariate, and influential observation sensitivity analyses are provided in the Supplemental Results (Tables S8 and S9; Figure S1).

## Discussion

In this study, we investigated whether the magnitude of interhemispheric ALPS differences provides prognostic information for Alzheimer disease conversion beyond bilateral mean ALPS and clinical and structural MRI measures. The key findings of this study were as follows: 1) greater absolute ALPS asymmetry was associated with AD conversion after adjustment for clinical variables, log-transformed WMH volume, BPF, and bilateral mean ALPS (adjusted HR per 1 SD, 1.26; P = .005), and its addition improved model fit (P = .01); 2) bilateral mean ALPS, hemisphere-specific ALPS, and the signed ALPS difference were not associated with conversion; and 3) adding absolute ALPS asymmetry did not consistently improve the cross-validated performance of survival models in predicting AD conversion. These findings support further evaluation of absolute ALPS asymmetry as a complementary marker of interhemispheric differences in periventricular diffusion.

The association between absolute ALPS asymmetry and AD conversion may be explained by a lateralized pathological process that bilateral averaging does not detect. In Rasmussen encephalitis, asymmetric ALPS reduction correlated with altered electroencephalographic aperiodic exponents within the atrophic cortex (28). In Parkinson disease, ALPS asymmetry differed according to the side of motor symptom onset (29). AD pathology also accumulates asymmetrically; baseline amyloid-β asymmetry predicted subsequent tau lateralization, and absolute tau laterality was associated with faster tau accumulation and cognitive decline after adjustment for bilateral tau burden (14,30). Across these conditions, the relatively preserved hemisphere did not compensate for the deficit on the affected side, and the magnitude of the left-right difference reflected the extent of lateralized pathology. Hemispheric differences in glymphatic transport could contribute to lateralized accumulation of AD pathology, with corresponding differences in periventricular diffusivity captured by absolute ALPS asymmetry.

Our results are in line with previous studies linking hemispheric asymmetry to prognosis. Greater ALPS asymmetry normalized by the bilateral mean was associated with poorer long-term seizure outcomes in MRI-negative focal epilepsy (17), and absolute tau asymmetry was associated with faster cognitive decline in AD (14,30). As in these tau PET studies, we tested the magnitude of asymmetry while adjusting for bilateral burden, and the association persisted after this adjustment and under both alternative asymmetry definitions. Our study extends previous ALPS studies by addressing the specific consideration that, unlike conditions examined in previous ALPS asymmetry studies (17,28,29,31), in which the affected side was known, the side of vulnerability in AD is not known a priori, necessitating a side-independent measure.

Consistent with a side-independent lateralized process in AD, neither hemisphere-specific ALPS nor the signed ALPS difference was associated with conversion. Because such a process can arise in either hemisphere, left- and right-dominant asymmetries offset each other in the signed difference, whereas the absolute difference quantifies the magnitude of imbalance regardless of side. Bilateral mean ALPS was also not associated with conversion, whereas previous studies reported associations with clinical progression (9,10). Our endpoint was the first AD diagnosis, our models were stratified by baseline diagnosis, and we included baseline MMSE, BPF, and log-transformed WMH volume as covariates.

The weak correlations with ventricular and cortical volume asymmetries suggest that absolute ALPS asymmetry captures information beyond the measured macroscopic structural differences. Absolute ALPS asymmetry was partly explained by association-fiber Dzz asymmetry, a component of the ALPS denominator, and the two measures carried overlapping prognostic information. Because Dzz in the association region is oriented perpendicular to both the dominant fiber direction and the medullary perivascular spaces, its asymmetry likely reflects asymmetric white matter microstructural injury, whereas the x-axis diffusivities also incorporate perivascular water diffusivity (32). Absolute ALPS asymmetry therefore may represent a structure-dependent periventricular diffusion phenotype that combines asymmetric perivascular water diffusivity and asymmetric white matter injury (11,32–35).

The improvement in Cox model fit reflects the contribution of absolute ALPS asymmetry across follow-up. The cross-validated 60-month AUC measures how well predicted risks rank participants who converted by that time above those who remained event-free. Adding a marker can change estimated risks while largely preserving their order. The gain in AUC can therefore be small when the reference model already discriminates well (36). In our study, reference AUCs exceeded 0.90, and adding asymmetry increased the 60-month AUC by 0.38 percentage points in both Cox regression and DeepSurv. Both confidence intervals included zero, leaving the magnitude of any discrimination gain uncertain at present.

Beyond its prognostic association, absolute ALPS asymmetry may also have clinical applications. Patients with similar bilateral mean ALPS differed in their estimated 5-year conversion risk according to the magnitude of interhemispheric differences. Absolute ALPS asymmetry could thus help identify patients who may benefit from closer cognitive follow-up or additional biomarker testing. Because it can be derived from existing diffusion MRI with an automated processing pipeline, it requires no additional acquisition and avoids manual ROI placement, which may reduce interobserver variability.

There are several limitations to our study. First, the retrospective ADNI design permits residual confounding and selection bias. Few conversions among CN participants limited diagnosis-specific interpretation, and clinical AD diagnoses do not establish molecular specificity. Second, diffusion MRI acquisition parameters were heterogeneous across sites, and the comprehensive technical adjustment excluded Philips examinations because of missing metadata. In addition, test-retest reliability could not be assessed in this retrospective cohort. Third, ALPS does not directly measure solute clearance, and its correspondence with intrathecal tracer measures remains uncertain (37). Fourth, incremental predictive value was not demonstrated, and external validation, incremental value beyond hippocampal measures, apolipoprotein E (APOE) genotype, and molecular biomarkers (38), and effects on clinical decisions were not assessed. Finally, death data were unavailable for competing-risk analysis, and secondary sensitivity analyses were not corrected for multiplicity.

In conclusion, greater absolute ALPS asymmetry was associated with subsequent AD conversion after adjustment for bilateral mean ALPS, clinical variables, log-transformed WMH volume, and brain parenchymal fraction. Considering interhemispheric heterogeneity in addition to the mean may extend ALPS-based prognostic assessment, and further investigation is warranted to establish its biological basis and clinical utility.

## Data Availability

Data collection and sharing for the Alzheimer's Disease Neuroimaging Initiative (ADNI) are funded by the National Institute on Aging (National Institutes of Health Grant U19AG024904). The grantee organization is the Northern California Institute for Research and Education. In the past, ADNI has also received funding from the National Institute of Biomedical Imaging and Bioengineering, the Canadian Institutes of Health Research, and private sector contributions through the Foundation for the National Institutes of Health (FNIH) including generous contributions from the following: AbbVie; Alzheimer's Association; Alzheimer's Drug Discovery Foundation; Araclon Biotech; BioClinica, Inc.; Biogen; Bristol-Myers Squibb Company; CereSpir, Inc.; Cogstate; Eisai Inc.; Elan Pharmaceuticals, Inc.; Eli Lilly and Company; EuroImmun; F. Hoffmann-La Roche Ltd and its affiliated company Genentech, Inc.; Fujirebio; GE Healthcare; IXICO Ltd.; Janssen Alzheimer Immunotherapy Research & Development, LLC.; Johnson & Johnson Pharmaceutical Research & Development LLC.; Lumosity; Lundbeck; Merck & Co., Inc.; Meso Scale Diagnostics, LLC.; NeuroRx Research; Neurotrack Technologies; Novartis Pharmaceuticals Corporation; Pfizer Inc.; Piramal Imaging; Servier; Takeda Pharmaceutical Company; and Transition Therapeutics. Individual deidentified participant data cannot be redistributed by the authors but are available to qualified investigators through the Alzheimer's Disease Neuroimaging Initiative, subject to its approval and data-use requirements. Study-specific code used for data preparation, statistical analyses, survival prediction, and figure generation is publicly available at https://github.com/MEDveritas/beyond-mean-alps. The automated DTI-ALPS image-processing pipeline used in this study is available at https://github.com/gbarisano/alps.

https://github.com/gbarisano/alps

## Abbreviations

AD: Alzheimer disease
ADNI: Alzheimer’s Disease Neuroimaging Initiative
ALPS: analysis along the perivascular space
AUC: area under the receiver operating characteristic curve
BPF: brain parenchymal fraction
CI: confidence interval
HR: hazard ratio
MCI: mild cognitive impairment
MMSE: Mini-Mental State Examination
WMH: white matter hyperintensity.

## Funding

This work was supported by the National Research Foundation of Korea (NRF) grant funded by the Korea government (MSIT) (NRF-2023R1A2C3003250, R.E.Y.). This study was supported by a grant from the Korea Health Technology R&D Project through the Korea Health Industry Development Institute (KHIDI), funded by the Ministry of Health & Welfare, Republic of Korea (RS-2023-00262321, R.E.Y.). This work was supported by the National Research Foundation of Korea (NRF) grant funded by the Korea government (MSIT) (IRIS RS-2026-25488643, R.E.Y., RS-2024-00358082, J.H.K.) and grant no. 0320230270 from the SNUH Research Fund (R.E.Y.). This work was supported by a grant of the Korea Health Technology R&D Project through the Korea Health Industry Development Institute (KHIDI), funded by the Ministry of Health & Welfare, Republic of Korea (RS-2025-02307233, S.H.C.).

### Acknowledgments

Data collection and sharing for the Alzheimer’s Disease Neuroimaging Initiative (ADNI) are funded by the National Institute on Aging (National Institutes of Health Grant U19AG024904). The grantee organization is the Northern California Institute for Research and Education. In the past, ADNI has also received funding from the National Institute of Biomedical Imaging and Bioengineering, the Canadian Institutes of Health Research, and private sector contributions through the Foundation for the National Institutes of Health (FNIH) including generous contributions from the following: AbbVie; Alzheimer’s Association; Alzheimer’s Drug Discovery Foundation; Araclon Biotech; BioClinica, Inc.; Biogen; Bristol-Myers Squibb Company; CereSpir, Inc.; Cogstate; Eisai Inc.; Elan Pharmaceuticals, Inc.; Eli Lilly and Company; EuroImmun; F. Hoffmann-La Roche Ltd and its affiliated company Genentech, Inc.; Fujirebio; GE Healthcare; IXICO Ltd.; Janssen Alzheimer Immunotherapy Research & Development, LLC.; Johnson & Johnson Pharmaceutical Research & Development LLC.; Lumosity; Lundbeck; Merck & Co., Inc.; Meso Scale Diagnostics, LLC.; NeuroRx Research; Neurotrack Technologies; Novartis Pharmaceuticals Corporation; Pfizer Inc.; Piramal Imaging; Servier; Takeda Pharmaceutical Company; and Transition Therapeutics.

## Data Sharing and Code Availability

Individual deidentified participant data cannot be redistributed by the authors but are available to qualified investigators through the Alzheimer’s Disease Neuroimaging Initiative, subject to its approval and data-use requirements. Study-specific code used for data preparation, statistical analyses, survival prediction, and figure generation is publicly available at https://github.com/MEDveritas/beyond-mean-alps. The automated DTI-ALPS image-processing pipeline used in this study is available at https://github.com/gbarisano/alps.

