## Supplemental Materials for "Greater Absolute DTI-ALPS Asymmetry Is Associated with Higher Risk of Conversion to Alzheimer Disease"

This supplement provides detailed methods, participant selection (Table S1), baseline diagnosis comparisons (Table S2), scanner models (Table S3), discrimination and prediction-error analyses (Tables S4-S5), directional, alternative definition, and tensor component analyses (Table S6), tensor component competition (Table S7), structural MRI sensitivity analyses (Table S8 and Figure S1), and technical sensitivity analyses (Table S9 and Figure S1).

### Supplemental Materials and Methods

#### 1 Data Source and Baseline Diffusion MRI

This retrospective longitudinal study used deidentified data from the Alzheimer's Disease Neuroimaging Initiative (ADNI). The ADNI protocol was approved by the institutional review board at each participating site, and all participants or their authorized representatives provided written informed consent. Data were linked across clinical, demographic, cognitive, structural MRI, and diffusion MRI tables by participant identifier. Screening, baseline, and screening-MRI visit codes were harmonized to a common baseline key where appropriate, and sentinel missing-value codes were recoded as missing before analysis.

Study baseline was defined as the date of each participant's first usable diffusion MRI scan. When more than one diffusion series was available on that date, the series with the greatest number of diffusion-encoding directions was selected. Baseline diagnostic status was assigned from the diagnostic assessment closest to the scan date within ±90 days. Diffusion MRI was obtained at the ADNI enrollment baseline visit in 912 participants (80.6%) and at a later ADNI visit in 219 (19.4%). Among the latter group, the median interval from ADNI enrollment to the baseline diffusion MRI scan was 72 months (IQR, 60–90 months).

#### 2 Clinical Variables and Outcome

Age was defined at study baseline. Sex and years of education were obtained from ADNI demographic records. Baseline Mini-Mental State Examination (MMSE) was defined as the assessment closest to the baseline diffusion MRI scan within the prespecified clinical window. When no value was available in that window, the closest MMSE assessment within ±365 days was used. The primary outcome was incident Alzheimer disease (AD), defined as the first site-adjudicated AD diagnosis after the diagnostic assessment used to define baseline. That visit date defined event time. Participants without conversion were right-censored at their last diagnostic assessment. Intermediate diagnostic fluctuations were not treated as events; no administrative censoring date was imposed.

**3 Structural MRI Measures**

Structural MRI variables were obtained from ADNI-processed tissue segmentation. Brain parenchymal fraction (BPF) was calculated as $\frac{\text{gray matter + normal−appearing white matter + WMH volume}}{\text{intracranial volume}}$. White matter hyperintensity (WMH) volume was log-transformed as ln(1 + WMH volume) because of right skew. BPF and log-transformed WMH volume were standardized before survival modeling. For structural asymmetry sensitivity analyses, lateral ventricular, cortical gray matter, and cortical white matter volumes were preferentially matched to the date of the baseline diffusion MRI scan and converted to structural asymmetry as $\frac{2|L-R|}{L+R}$.

Structural MRI covariates were derived from the University of California, Davis four-tissue segmentation pipeline. Gray matter, white matter, and cerebrospinal fluid were segmented from T1-weighted images using an expectation-maximization/Markov-random-field framework; WMH were identified using a dedicated Bayesian segmentation of coregistered T1-weighted and FLAIR images. In the final four-tissue segmentation, voxels classified as WMH replaced their corresponding labels in the three-tissue segmentation, yielding mutually exclusive gray matter, white matter, cerebrospinal fluid (CSF), and WMH classes. Volumes were reported in native-space cubic centimeters.

**4 Diffusion MRI Acquisition and Preprocessing**

All diffusion inputs used for ALPS processing were recorded at 3-T on Siemens (n = 600), GE Healthcare (n = 379), or Philips (n = 152) systems. Gradient files documented 857 single-shell and 274 multi-shell acquisitions when diffusion shells were defined using b ≥ 50 s/mm². Scanner model names were available for all participants; model-specific counts are provided in Table S3. Single-shell configurations were b =

1000 s/mm² (n = 798), b = 1300 s/mm² (n = 9), and b = 2000 s/mm² (n = 50). Multi-shell configurations were b = 500/1000/2000 s/mm² (n = 264), b = 500/1000/1950/2000 s/mm² (n = 9), and b = 1000/2000 s/mm² (n = 1). The selected inputs contained 30–126 diffusion-weighted volumes (median, 54). Native stored in-plane pixel spacing ranged from 0.91 to 3.0 mm (median, 2.0 mm), section thickness from 2.0 to 4.0 mm (median, 2.0 mm), repetition time from 3,067 to 17,523 ms (median, 7,800 ms), and echo time from 47.7 to 105.0 ms (median, 71.0 ms). Because stored in-plane pixel spacing may include scanner-side reconstruction interpolation, these values were considered stored image spacing rather than a uniform measure of intrinsic acquisition sampling resolution.

Automated DTI-ALPS processing was performed using a publicly available pipeline with FSL 6.0 and MRtrix3 3.0.8 (20,21). Diffusion-weighted images underwent denoising and Gibbs-ringing correction, followed by eddy-current and head-motion correction with FSL eddy and diffusion-tensor fitting. Fractional anisotropy (FA) maps were affinely registered to the JHU-ICBM-FA 1-mm template using FLIRT with 12 degrees of freedom, the correlation-ratio cost function with 256 bins, and rotation searches from −90° to 90° along each axis. The resulting affine transformation was applied to the diffusion-tensor volumes using FSL applywarp (20).

**5 ALPS Calculation and Quality Control**

Four spherical regions of interest (ROIs) with a 2.5-mm radius sampled the bilateral superior corona radiata (projection fibers) and superior longitudinal fasciculus (association fibers) adjacent to the bodies of the lateral ventricles. The ROI centers in JHU-ICBM-FA 1-mm template-space coordinates were (−26, −16, 27) and (26, −16, 27) mm for the left and right projection regions and (−38, −16, 27) and (38, −16, 27) mm for the left and right association regions, respectively. For each hemisphere, ALPS was calculated as $\frac{D_{\mathrm{xx,projection}} + D_{\mathrm{xx,association}}}{D_{\mathrm{yy,projection}} + D_{\mathrm{zz,association}}}$. Bilateral mean ALPS was $\frac{\mathrm{ALPS_L} + \mathrm{ALPS_R}}{2}$.

The primary exposure was absolute ALPS asymmetry, defined as the raw absolute left-right difference in the ALPS index, $|\mathrm{ALPS_L} - \mathrm{ALPS_R}|$. Scale-independent sensitivity definitions were mean-normalized relative asymmetry, $\frac{2|\mathrm{ALPS_L} - \mathrm{ALPS_R}|}{\mathrm{ALPS_L} + \mathrm{ALPS_R}}$, and the absolute log ratio, $\left|\log\left(\frac{\mathrm{ALPS_L}}{\mathrm{ALPS_R}}\right)\right|$. The signed

ALPS difference (L−R) was evaluated separately to distinguish asymmetry magnitude from hemispheric direction. Each continuous imaging measure was standardized before modeling.

Sixteen participants had ALPS processing failures: nine had invalid diffusion reconstructions (two missing gradient tables, six b0-only acquisitions, and one examination with incomplete brain coverage), and seven had unrecoverable eddy-current correction failures. After successful processing, eight regional diffusivity components and left, right, and bilateral mean ALPS were screened at ±3 SD. Fifty-eight flagged examinations were reviewed by one radiologist on template-space FA images with all four regions overlaid. Review assessed periventricular white matter placement, left-right symmetry, and image integrity. Statistical flagging alone was not an exclusion criterion; visual review prompted no additional exclusions.

### 6 Primary Cox Model and Adjusted Absolute Risk

For the Cox models specified in the main text, continuous predictors were standardized at the full-cohort mean and scaled using the population SD. Sex was coded 0 for male and 1 for female. The complete-case sample contained 1,131 participants and 115 AD conversions. Schoenfeld residual tests used rank-transformed event times. Robust standard errors were calculated without an additional cluster variable. Cox analyses used Python 3.12.4 and lifelines 0.30.0.

#### 6.1 Adjusted Absolute Risk

To complement relative hazard estimates with clinically interpretable absolute probabilities, adjusted AD conversion risks were estimated at 36 and 60 months by marginal standardization from the primary Cox model. Each participant's observed covariates and diagnosis stratum were retained while absolute ALPS asymmetry was set to the cohort mean (0 SD) or 1 SD above the mean; predicted conversion probabilities were averaged over the analytic cohort. Month t was converted to t × 365.25/12 days before survival prediction; conversion probability was 1 minus mean predicted survival. The 0-SD and +1 SD settings corresponded to raw asymmetry values of approximately 0.110 and 0.202, respectively. The absolute risk difference was calculated by subtracting the unrounded 0-SD probability from the unrounded +1 SD probability. Percentile 95% CIs were obtained from 500 participant-level bootstrap replicates.

#### 6.2 Continuous Association Display

An exploratory restricted cubic spline Cox model included 1,131 participants and 115 AD conversions. The exposure was standardized using the sample SD; three knots were placed at the 10th, 50th, and 90th percentiles of absolute ALPS asymmetry (approximately 0.014, 0.088, and 0.229). The model had two exposure degrees of freedom (one linear and one nonlinear) and adjusted for sex, age, education, baseline MMSE, log-transformed WMH volume, BPF, and bilateral mean ALPS, with baseline-diagnosis stratification and follow-up in days.

Likelihood-ratio tests compared the spline model with a model omitting asymmetry (2 df; $\chi^2 = 6.73$; P = .035) and with a linear asymmetry model using the same spline-analysis covariates (1 df; $\chi^2 = 0.30$; P = .58). Relative hazards were referenced to median raw asymmetry (0.088). Pointwise 95% CIs were computed as $\exp\left(\Delta b\beta \pm 1.96\sqrt{\Delta b V \Delta b^{\mathrm{T}}}\right)$, where Δb is the spline-basis contrast against the median and V is the estimated covariance matrix of the fitted spline coefficients. The fit used all complete observations; 240 prediction coordinates spanned the 1st-99th percentiles (0.008–0.287 across the 5th-95th percentiles). The spline model used the same ln(1 + WMH volume) transformation, day-based follow-up, covariates, and baseline-diagnosis stratification as the primary linear Cox model; only the exposure functional form differed.

### 7 Sensitivity Analyses

#### 7.1 Directional Component, Alternative Definition, Tensor Component, and Outlier Sensitivity Analyses

Left ALPS, right ALPS, and bilateral mean ALPS were tested separately with adjustment for age, sex, education, baseline MMSE, log-transformed WMH volume, and BPF, stratified by baseline diagnosis. Models of the signed ALPS difference and absolute ALPS asymmetry additionally adjusted for bilateral mean ALPS (primary Cox model). Thus, the single-marker bilateral mean ALPS estimate differs from its coefficient in the primary Cox model. The two alternative asymmetry definitions and the absolute left-right asymmetry of each directional diffusivity component (projection-fiber Dxx, association-fiber Dxx, projection-fiber Dyy, and association-fiber Dzz) were evaluated separately, with additional adjustment for bilateral mean ALPS. Joint models tested whether the primary signal was reducible to a single component, particularly association-fiber Dzz asymmetry. Two-sided Spearman correlations quantified the overlap between absolute ALPS asymmetry and each component's absolute left-right asymmetry. In the analysis of absolute ALPS asymmetry and association-fiber Dzz asymmetry, both exposures were raw absolute left-right differences, standardized with full-cohort means and SDs. Four models compared the reference covariates alone, each exposure separately, and both together on the same 1,131 participants. Nested LR tests distinguished joint two-degree-of-freedom contribution from each exposure's residual one-degree-of-freedom contribution. We refitted the joint Cox model in 300 participant-level bootstrap resamples using the original standardized values and model-based standard errors. Wald inference in the observed sample used robust sandwich standard errors.

Robustness to extreme and influential observations was assessed by excluding the upper 1%, 2.5%, and 5% of the absolute ALPS asymmetry distribution and by removing the 1, 5, or 10 observations with the largest delta-beta influence on the absolute ALPS asymmetry coefficient. Full-cohort standardization was retained after exclusion. Sensitivity analyses used nominal two-sided P values.

#### 7.2 Technical and Structural Analyses

Scanner and protocol sensitivity covariates included scanner manufacturer, number of diffusion-encoding directions, section thickness, repetition time, and echo time. Within-scan motion was summarized primarily by mean relative root-mean-square (RMS) displacement from the second column of the FSL eddy movement-RMS output. Restricted relative RMS, P95 relative RMS, and outlier-slice fraction were evaluated separately as sensitivity measures. Mean and 95th-percentile relative RMS were measured in millimeters. Outlier-slice fraction was the number of nonzero entries divided by the total number of entries in the eddy outlier map. Motion metrics were obtained from the FSL eddy outputs for the baseline diffusion MRI scans.

Registration-derived head orientation was estimated by rigid six-degree-of-freedom registration of native FA to the JHU-ICBM-FA template and represented as pitch, yaw, and roll. DICOM acquisition-plane orientation was derived separately from ImageOrientationPatient direction cosines: normalized row and column vectors were crossed to obtain the slice normal, from which two plane-tilt angles were calculated.

Head orientation and acquisition-plane orientation were therefore treated as distinct technical domains. FLIRT used six degrees of freedom and correlation-ratio cost. FSL avscale extracted x-, y-, and z-axis rotations, which were converted from radians to degrees and recorded as roll, pitch, and yaw, respectively.

For each technical block, the expanded model and its reference model were fitted on the same complete-case subset. A comprehensive model jointly included scanner/protocol, motion, head orientation, and acquisition-plane variables. The full primary cohort included 1,131 participants and 115 events. Same-day structural adjustment included 1,097 participants and 110 events, and comprehensive technical adjustment included 936 participants and 105 events. Reductions reflected availability of the required measurements, and the comparison samples were specific to each analysis. Same-sample refitting does not establish that complete-case selection is unbiased. The comprehensive technical sample excluded 195 participants, including 10 converters, because diffusion-direction counts were unavailable. Technical Cox models used the same ln(1 + WMH volume) transformation as the primary Cox model. Continuous technical-model variables were standardized using their available full-cohort values, without restandardization after common-sample selection. Nonvarying manufacturer indicators were omitted from Cox fitting.

Structural asymmetry analyses used ADNI-provided UCSFFSX7 (UCSF cross-sectional FreeSurfer 7.x) measurements. Left/right lateral ventricular, cortical gray matter, and cortical white matter volumes were ST37SV/ST96SV, ST147SV/ST148SV, and ST150SV/ST151SV, respectively. Asymmetry was $\frac{2|L-R|}{L+R}$. Same-day records were prioritized; duplicate records on the same date were ranked by processing status, QC rank, and structural-data completeness. Spearman correlations used pairwise-complete observations. Cox models used a common same-day sample of 1,097 participants with 110 events and evaluated separate and combined additions of ventricular burden and structural asymmetries; not every adjacent model was nested. Basic continuous covariates and ALPS measures retained full-cohort standardization, whereas structural measures were standardized using the common-sample mean and SD. Models used ln(1 + WMH volume) and day-derived follow-up. An additional analysis selected the closest structural examination within ±365 days, choosing the earlier date when equally distant; its final model included 1,101 participants and 111 events. This temporal-matching sensitivity reported the fitted asymmetry association without a separate paired reduced-model LR test.

### 8 Incremental Survival Prediction

The prediction analyses assessed whether absolute ALPS asymmetry improved prediction of AD conversion beyond clinical and structural predictors and bilateral mean ALPS. Conventional Cox regression and DeepSurv are the focus of the main-text comparison. CoxTime and DeepHitSingle were evaluated on the same outer splits and are reported in Tables S4-S5. Together, these analyses characterized the prognostic contribution of absolute ALPS asymmetry across discrimination and prediction error.

Reference predictors comprised age, sex, education, baseline diagnosis, baseline Mini-Mental State Examination score, brain parenchymal fraction (BPF), log-transformed white matter hyperintensity (WMH) volume, and bilateral mean ALPS. Extended models additionally included absolute ALPS asymmetry, defined as $|\mathrm{ALPS_L} - \mathrm{ALPS_R}|$, without normalization by bilateral mean ALPS. Both predictor sets were evaluated in the same complete-case cohort of 1,131 participants, including 115 AD conversions.

#### 8.1 Cross-Validation and Model Training

Models used five repetitions of five-fold cross-validation, with participant splits stratified jointly by baseline diagnosis and AD conversion status. Within each repetition, identical outer folds were used across models and predictor sets. For the neural models, within each outer-training set, one of five stratified inner folds was reserved for early stopping. Standardization parameters were fitted using the remaining inner-training participants and applied to validation and held-out test participants. Survival-time transformations were also fitted using training data only. The conventional Cox model used the outer training set, standardized continuous predictors within that set, and estimated separate baseline hazards for CN and MCI. For this model baseline diagnosis was a stratification variable.

All architectures used two hidden layers with 16 and 8 units and dropout of 0.15. Adam optimization used a learning rate of 0.001, a maximum of 256 epochs, and early-stopping patience of 25 epochs. DeepSurv used full training-set batches; CoxTime and DeepHitSingle used batches of 256. DeepHitSingle used 24 duration bins, alpha of 0.20, and sigma of 0.10. Three training runs, using seed offsets of 111, 222, and 333, were averaged within each outer fold. Each participant therefore contributed one held-out prediction per model and predictor set in each repetition. Analyses used Python 3.12.4 and pycox 0.3.0. Batch normalization was disabled. DeepHitSingle survival curves used a 10-fold interpolation factor before interpolation to the evaluation horizons. To ensure reproducibility, data partitioning and stochastic model training were controlled using fixed random seeds (exact seed schedules are provided in the public code repository). Outer-split seeds were 20260828 + 1009 × repetition; inner-split seeds were 20260828 + 10000 × repetition + fold, and training seeds additionally included 111, 222, or 333, with repetition and fold indexed from zero.

#### 8.2 Discrimination and Prediction Error

Follow-up in months was calculated from elapsed days divided by 365.25/12. Predicted AD conversion probability was one minus predicted survival. Cumulative/dynamic area under the receiver operating characteristic curve (AUC) and inverse-probability-of-censoring-weighted (IPCW) Brier scores were evaluated at 12, 36, and 60 months. Cases converted by the horizon; dynamic controls were observed beyond it. Participants censored at or before the horizon contributed neither as cases nor as controls to AUC estimation.

AUC used the saved fold-specific case and control censoring weights. Case-control concordance was 1 when predicted risk was higher in the case, 0 when lower, and 0.5 for a tie. Contributions were weighted by the product of the case and control weights and normalized by total pair weight. AUC and prediction-error metrics were calculated within each repetition and then averaged across five repetitions, rather than calculated from predictions averaged across repetitions. For each outer fold, the censoring distribution G was estimated from all outer-training participants using Kaplan-Meier with censoring as the event. Case weights were $\frac{1}{G(T^-)}$ at the observed conversion time T; control weights were $\frac{1}{G(t)}$ at the horizon t. A horizon was unsupported when it reached or exceeded the maximum outer-training follow-up or the required censoring survival was below 0.05.

The integrated Brier score (IBS) used trapezoidal integration on a 3-month grid from 12 to 60 months, divided by the 48-month interval. Positive extended-minus-reference ΔAUC and negative Brier-score or IBS differences favored the extended model. ΔAUC is reported in percentage points; Brier scores, IBS, and their differences are multiplied by 1,000. The censoring-survival threshold was 0.05; horizons with fewer than 20 cases or controls were flagged as having limited information. At 36 months, there were 59 cases, 571 controls, and 501 earlier-censored participants; corresponding 60-month counts were 90, 388, and 653. IBS was also flagged because its grid included the 12-month horizon with only 13 cases.

#### 8.3 Bootstrap Inference and Multiplicity

Paired extended-minus-reference differences were evaluated using 300 participant-level bootstrap samples. Resampling preserved each participant's paired predictions and records across repetitions. Prediction models and censoring weights remained fixed. Point estimates were calculated from the observed data, and percentile 95% confidence intervals were obtained from the bootstrap distribution. These intervals quantify sampling uncertainty conditional on the fitted out-of-fold predictions and censoring weights and do not incorporate uncertainty from refitting either component. At least 90% of requested bootstrap samples were required to yield valid estimates. Approximate two-sided bootstrap P values were twice the smaller tail proportion relative to zero, with one added to both the tail count and the number of valid replicates, and were capped at 1. Finite-resample tail estimates and interpolated percentile limits can differ at the significance boundary. Conventional Cox AUC contrasts used a separate set of 10,000 participant-level paired bootstrap resamples with the same fixed-prediction and fixed-weight evaluation.

The original 60-month ΔAUC family comprised three neural-model contrasts (Holm adjustment). The secondary neural family comprised 36-month ΔAUC, 36- and 60-month Brier-score contrasts, and IBS contrasts (Benjamini-Hochberg adjustment). Conventional Cox AUC P values were nominal (Tables S4-S5).

### Supplemental Results

#### Sensitivity Analyses for Influential Observations, Structural Asymmetry, and Technical Covariates

Absolute ALPS asymmetry showed little correlation with ventricular (n = 1,099, Spearman ρ = 0.02, P = .46), cortical gray matter (n = 1,100, ρ = 0.01, P = .82), or cortical white matter asymmetry (n = 1,097, ρ = −0.05, P = .10). In the same-day structural sample of 1,097 participants, the adjusted HR after structural covariate adjustment was 1.22 (95% CI: 1.04, 1.44; P = .02) (Table S8).

After comprehensive technical adjustment in 936 participants, the HR was 1.23 (95% CI: 1.04, 1.46; P = .02) (Table S9, Figure S1). Estimates were similar after influential observations were removed. Excluding the upper 5% of absolute asymmetry values reduced precision.

### Supplemental Tables

#### Table S1. Participant Selection

| Step | Exclusion criterion | Excluded, n | Remaining, n |
|---|---|---|---|
| 0 | Diffusion-MRI eligible with ≥1 dated diagnostic record | — | 1,846 |
| 1 | No diagnostic assessment within ±90 days of the baseline diffusion MRI scan | 323 | 1,523 |
| 2 | AD diagnosis at study baseline | 178 | 1,345 |
| 3 | No diagnostic assessment after baseline | 141 | 1,204 |
| 4 | Baseline MMSE neither observed nor recoverable | 2 | 1,202 |
| 5 | Fewer than 30 diffusion-encoding directions | 8 | 1,194 |
| 6 | Required structural imaging volumes unavailable | 47 | 1,147 |
| 7 | ALPS processing failures | 16 | 1,131 |
| 7a | Invalid diffusion reconstruction | 9 | — |
| | Missing gradient table | 2 | — |
| | b0-only acquisition | 6 | — |
| | Incomplete brain coverage | 1 | — |
| 7b | Unrecoverable eddy-current correction failure | 7 | — |

Exclusions were sequential. Rows 7a and 7b subdivide the 16 processing failures in step 7 and are not additional exclusions. The three reconstruction subcategories sum to nine. The final cohort comprised 669 cognitively normal participants and 462 with mild cognitive impairment. AD = Alzheimer disease; ALPS = analysis along the perivascular space; MMSE = Mini-Mental State Examination.

**Table S2. Baseline Participant Characteristics by Baseline Diagnosis**

| Characteristic | Overall (n = 1,131) | CN (n = 669) | MCI (n = 462) | P |
|---|---|---|---|---|
| Age, years | 72.1 ± 7.7 | 71.1 ± 7.6 | 73.5 ± 7.7 | < .001 |
| Women, n (%) | 607 (53.7) | 402 (60.1) | 205 (44.4) | < .001 |
| Education, years | 16.4 ± 2.5 | 16.6 ± 2.4 | 16.1 ± 2.6 | < .001 |
| Baseline MMSE | 28.4 ± 1.7 | 29.0 ± 1.2 | 27.5 ± 2.0 | < .001 |
| WMH volume, mL | 1.95 (0.61–5.45) | 1.5 (0.46–4.0) | 2.8 (0.96–7.3) | < .001 |
| BPF, % | 75.3 ± 2.9 | 75.9 ± 2.6 | 74.5 ± 3.0 | < .001 |
| Bilateral mean ALPS | 1.26 ± 0.19 | 1.28 ± 0.19 | 1.23 ± 0.20 | < .001 |
| Absolute ALPS asymmetry | 0.11 ± 0.09 | 0.11 ± 0.09 | 0.11 ± 0.10 | .67 |

Continuous data are mean ± SD or median (IQR). P values compare the CN and MCI groups and were calculated using two-sided Welch t tests for variables reported as mean ± SD, Mann-Whitney U tests for variables reported as median (IQR), and a Pearson $\chi^2$ test for sex. BPF is expressed as a percentage. ALPS = analysis along the perivascular space; BPF = brain parenchymal fraction; CN = cognitively normal; IQR = interquartile range; MCI = mild cognitive impairment; MMSE = Mini-Mental State Examination; SD = standard deviation; WMH = white matter hyperintensity.

**Table S3. Scanner Models at Baseline**

| Manufacturer | Scanner model | n |
|---|---|---|
| Siemens | Prisma fit | 312 |
| Siemens | Prisma | 98 |
| Siemens | Skyra | 91 |
| Siemens | Verio | 53 |
| Siemens | TrioTim | 35 |
| Siemens | Vida | 6 |
| Siemens | Biograph mMR | 2 |
| Siemens | Cima.X | 2 |
| Siemens | Skyra fit | 1 |
| GE Healthcare | Discovery MR750 | 186 |
| GE Healthcare | SIGNA HDxt | 83 |
| GE Healthcare | SIGNA Premier | 44 |
| GE Healthcare | Discovery MR750w | 42 |
| GE Healthcare | SIGNA UHP | 17 |
| GE Healthcare | SIGNA HDx | 7 |
| Philips | Ingenia | 61 |
| Philips | Achieva | 43 |
| Philips | Achieva dStream | 38 |
| Philips | Intera | 5 |
| Philips | MR 7700 | 4 |
| Philips | Ingenia Elition X | 1 |
| Total | | 1,131 |

Counts refer to the selected baseline diffusion MRI scan for each participant (N = 1,131). Manufacturer and model names were harmonized for capitalization, underscores, and the MAGNETOM prefix. Prisma_fit and MAGNETOM Prisma Fit, Prisma and MAGNETOM Prisma, Skyra and MAGNETOM Skyra, and DISCOVERY MR750w and Discovery MR750w were combined, respectively. Prisma fit and Skyra fit were retained as separate entries from Prisma and Skyra.

**Table S4. Full Cohort Incremental Discrimination After Adding Absolute ALPS Asymmetry**

| Horizon, mo | Model | Reference AUC | Extended AUC | ΔAUC, percentage points (95% CI) | Adjusted P |
|---|---|---|---|---|---|
| 36 | DeepSurv | 0.9055 | 0.9059 | +0.05 (−0.72, +0.83) | .93 |
| 36 | CoxTime | 0.8890 | 0.8822 | −0.68 (−1.99, +0.57) | .55 |
| 36 | DeepHitSingle | 0.9038 | 0.9072 | +0.34 (−0.57, +1.24) | .58 |
| 36 | Conventional Cox | 0.9084 | 0.9093 | +0.09 (−0.57, +0.77) | — |
| 60 | DeepSurv | 0.9014 | 0.9052 | +0.38 (−0.55, +1.54) | > .99 |
| 60 | CoxTime | 0.8903 | 0.8870 | −0.33 (−1.96, +1.03) | > .99 |
| 60 | DeepHitSingle | 0.8969 | 0.9008 | +0.39 (−0.47, +1.29) | > .99 |
| 60 | Conventional Cox | 0.9141 | 0.9178 | +0.38 (−0.32, +1.13) | — |

Reference covariates included age, sex, education, baseline MMSE, BPF, log-transformed WMH volume, and bilateral mean ALPS. Diagnosis was a predictor for the neural models and a stratum for conventional Cox. Extended models added absolute ALPS asymmetry. Repetition-specific AUCs were averaged across five repetitions. Paired percentile 95% CIs used fixed out-of-fold predictions and censoring weights, with 300 bootstrap resamples for neural models and 10,000 for conventional Cox. Positive ΔAUC favors the extended model. Neural-model P values used Holm adjustment at 60 months and Benjamini-Hochberg adjustment at 36 months. Conventional Cox nominal P values were .78 and .31 at 36 and 60 months, respectively. At 36 months there were 59 cases, 571 dynamic controls, and 501 earlier-censored participants. At 60 months, the corresponding counts were 90, 388, and 653. ALPS = analysis along the perivascular space; AUC = area under the receiver operating characteristic curve; BPF = brain parenchymal fraction; CI = confidence interval; MMSE = Mini-Mental State Examination; WMH = white matter hyperintensity.

**Table S5. Incremental Prediction Error After Adding Absolute ALPS Asymmetry**

| Horizon, mo | Model | Metric | Difference ×1,000 (95% CI) | Adjusted P |
|---|---|---|---|---|
| 36 | DeepSurv | Brier | −0.564 (−1.773, +0.544) | .55 |
| 36 | CoxTime | Brier | +0.976 (−0.359, +2.438) | .48 |
| 36 | DeepHitSingle | Brier | −0.465 (−1.165, +0.157) | .48 |
| 60 | DeepSurv | Brier | +0.296 (−1.645, +2.233) | .86 |
| 60 | CoxTime | Brier | +2.292 (+0.046, +4.442) | .35 |
| 60 | DeepHitSingle | Brier | −0.613 (−1.731, +0.348) | .48 |
| 12–60 | DeepSurv | IBS | −0.223 (−1.143, +0.703) | .82 |
| 12–60 | CoxTime | IBS | +0.997 (−0.092, +2.211) | .35 |
| 12–60 | DeepHitSingle | IBS | −0.508 (−1.089, +0.005) | .35 |

Differences are extended minus reference and are multiplied by 1,000; negative differences favor the extended model. Brier scores use IPCW, and IBS integrates Brier scores over 12–60 months. Adjusted P values use the secondary Benjamini-Hochberg family. The 60-month CoxTime Brier increase had an unadjusted approximate P = .05 but adjusted P = .35. All IBS-difference intervals included zero; IBS was flagged as low information because its grid includes sparse early events. ALPS = analysis along the perivascular space; CI = confidence interval; IBS = integrated Brier score; IPCW = inverse probability of censoring weighting.

**Table S6. Directional, Alternative Definition, and Tensor Component Cox Analyses**

| Exposure Measure | HR per 1 SD (95% CI) | Wald P | LR P |
|---|---|---|---|
| **Hemispheric and bilateral mean ALPS** | | | |
| Left ALPS | 0.98 (0.80, 1.21) | .87 | .87 |
| Right ALPS | 0.97 (0.79, 1.20) | .79 | .78 |
| Bilateral mean ALPS (model without absolute ALPS asymmetry) | 0.98 (0.80, 1.20) | .81 | .82 |
| **Primary asymmetry vs signed difference** | | | |
| Signed ALPS difference (L-R) | 1.02 (0.82, 1.25) | .89 | .88 |
| Absolute ALPS asymmetry (primary Cox model) | 1.26 (1.07, 1.48) | .005 | .01 |
| **Alternative scale-independent asymmetry definitions** | | | |
| Mean-normalized relative asymmetry | 1.22 (1.05, 1.42) | .01 | .02 |
| Absolute log ratio | 1.22 (1.05, 1.42) | .01 | .02 |
| **Absolute asymmetry of individual tensor components** | | | |
| Projection-fiber Dxx asymmetry | 0.95 (0.83, 1.09) | .48 | .56 |
| Association-fiber Dxx asymmetry | 1.19 (1.01, 1.39) | .03 | .04 |
| Projection-fiber Dyy asymmetry | 1.07 (0.92, 1.24) | .36 | .42 |
| Association-fiber Dzz asymmetry | 1.19 (1.07, 1.32) | .001 | .01 |

All models were fitted in the full analytic cohort (n = 1,131; 115 AD conversions) and adjusted for age, sex, education, baseline MMSE, log-transformed WMH volume, and BPF, with stratification by baseline diagnosis. The left, right, and bilateral mean ALPS estimates came from separate models. The signed difference, absolute ALPS asymmetry, the two alternative asymmetry definitions, and each of the four tensor component asymmetries also adjusted for bilateral mean ALPS. The bilateral mean ALPS row shows the single-marker model. LR P compares each model with its corresponding reduced model on the same sample. AD = Alzheimer disease; ALPS = analysis along the perivascular space; BPF = brain parenchymal fraction; CI = confidence interval; Dxx = x-axis diffusivity; Dyy = y-axis diffusivity; Dzz = z-axis diffusivity; HR = hazard ratio; LR = likelihood-ratio; MMSE = Mini-Mental State Examination; SD = standard deviation; WMH = white matter hyperintensity.

**Table S7. Competition Between Absolute ALPS Asymmetry and Association-Fiber Dzz Asymmetry**

| Model | Marker | n | HR per 1 SD (95% CI) | Wald P |
|---|---|---|---|---|
| Absolute ALPS asymmetry only | Absolute ALPS asymmetry | 1,131 | 1.26 (1.07, 1.48) | .005 |
| Association-fiber Dzz asymmetry only | Association-fiber Dzz asymmetry | 1,131 | 1.19 (1.07, 1.32) | .001 |
| Joint model | Absolute ALPS asymmetry | 1,131 | 1.19 (0.99, 1.43) | .07 |
| Joint model | Association-fiber Dzz asymmetry | 1,131 | 1.14 (1.00, 1.29) | .05 |

Association-fiber Dzz asymmetry is the absolute left-right difference in Dzz within the association-fiber ROIs in the ALPS coordinate frame. In 300 participant-level bootstrap replicates of the joint model, median HRs were 1.19 for absolute ALPS asymmetry and 1.15 for association-fiber Dzz asymmetry. All 300 bootstrap replicates succeeded, and original full-cohort standardization was retained in each resample. All models included 1,131 participants and 115 AD conversions. ALPS = analysis along the perivascular space; CI = confidence interval; Dzz = z-axis diffusivity; HR = hazard ratio; SD = standard deviation.

**Table S8. Cox Sensitivity Analysis with Structural MRI Covariates**

| Model Analysis | n | Events | HR per 1 SD (95% CI) | Wald P | β Attenuation |
|---|---|---|---|---|---|
| **Same-day structural MRI cohort** | | | | | |
| Reference model | 1,097 | 110 | 1.26 (1.08, 1.48) | .004 | Reference |
| + Ventricular volume and asymmetry | 1,097 | 110 | 1.24 (1.05, 1.46) | .01 | 8.1% |
| + Ventricular volume and all structural asymmetries | 1,097 | 110 | 1.22 (1.04, 1.44) | .02 | 14.5% |
| **Temporal-matching sensitivity (±365 days)** | | | | | |
| + Ventricular volume and all structural asymmetries | 1,101 | 111 | 1.22 (1.03, 1.43) | .02 | — |

HRs are adjusted hazard ratios for absolute ALPS asymmetry. Same-day common-sample models included 1,097 participants and 110 events. Structural covariates comprised total lateral ventricular volume and left-right asymmetries of lateral ventricular, cortical gray matter, and cortical white matter volumes. Basic covariates retained full-cohort scaling, and structural covariates used common-sample scaling. The ±365-day analysis included 1,101 participants and 111 events. ALPS = analysis along the perivascular space; CI = confidence interval; HR = hazard ratio; SD = standard deviation.

**Table S9. Technical Covariate Adjustment for Absolute ALPS Asymmetry: Cox Sensitivity Analyses**

| Technical block | n | Events | Reference HR (95% CI) | Adjusted HR (95% CI) | Wald P | β attenuation |
|---|---|---|---|---|---|---|
| Scanner/protocol | 936 | 105 | 1.24 (1.05, 1.47) | 1.20 (1.01, 1.41) | .035 | 18.04% |
| Within-scan motion | 1,131 | 115 | 1.26 (1.07, 1.48) | 1.27 (1.08, 1.49) | .004 | −2.95% |
| Head orientation | 1,131 | 115 | 1.26 (1.07, 1.48) | 1.28 (1.09, 1.50) | .002 | −6.81% |
| Acquisition-plane orientation | 1,131 | 115 | 1.26 (1.07, 1.48) | 1.27 (1.08, 1.49) | .004 | −2.80% |
| Motion + head orientation | 1,131 | 115 | 1.26 (1.07, 1.48) | 1.29 (1.10, 1.51) | .002 | −9.57% |
| All technical except acquisition plane | 936 | 105 | 1.24 (1.05, 1.47) | 1.25 (1.05, 1.48) | .01 | −0.83% |
| All technical covariates | 936 | 105 | 1.24 (1.05, 1.47) | 1.23 (1.04, 1.46) | .02 | 4.71% |

Reference and expanded models were fitted on the same complete-case subset for each row. Negative attenuation indicates a slightly larger asymmetry coefficient after adjustment. Sample reductions reflect missing required technical measurements; same-sample comparisons do not establish absence of selection bias. Technical-model reference and expanded fits used ln(1 + WMH volume), matching the primary Cox model in Table 2. The reduction to 936 participants was attributable to missing diffusion-direction counts in 195 participants, including 10 converters. The 936-participant technical complete-case sample comprised 557 Siemens, 379 GE Healthcare, and no Philips examinations. Among the 195 participants without diffusion-direction-count metadata, the corresponding counts were 43, 0, and 152, respectively. Because no Philips examinations were represented in the complete-case sample, manufacturer-related sensitivity analyses could only evaluate variation between Siemens and GE Healthcare and therefore should not be interpreted as demonstrating robustness across all three manufacturers. Wald P reports the asymmetry coefficient's Wald P after covariate adjustment, not a multiplicity-adjusted P value. ALPS = analysis along the perivascular space; CI = confidence interval; HR = hazard ratio; WMH = white matter hyperintensity.

### Supplemental Figure

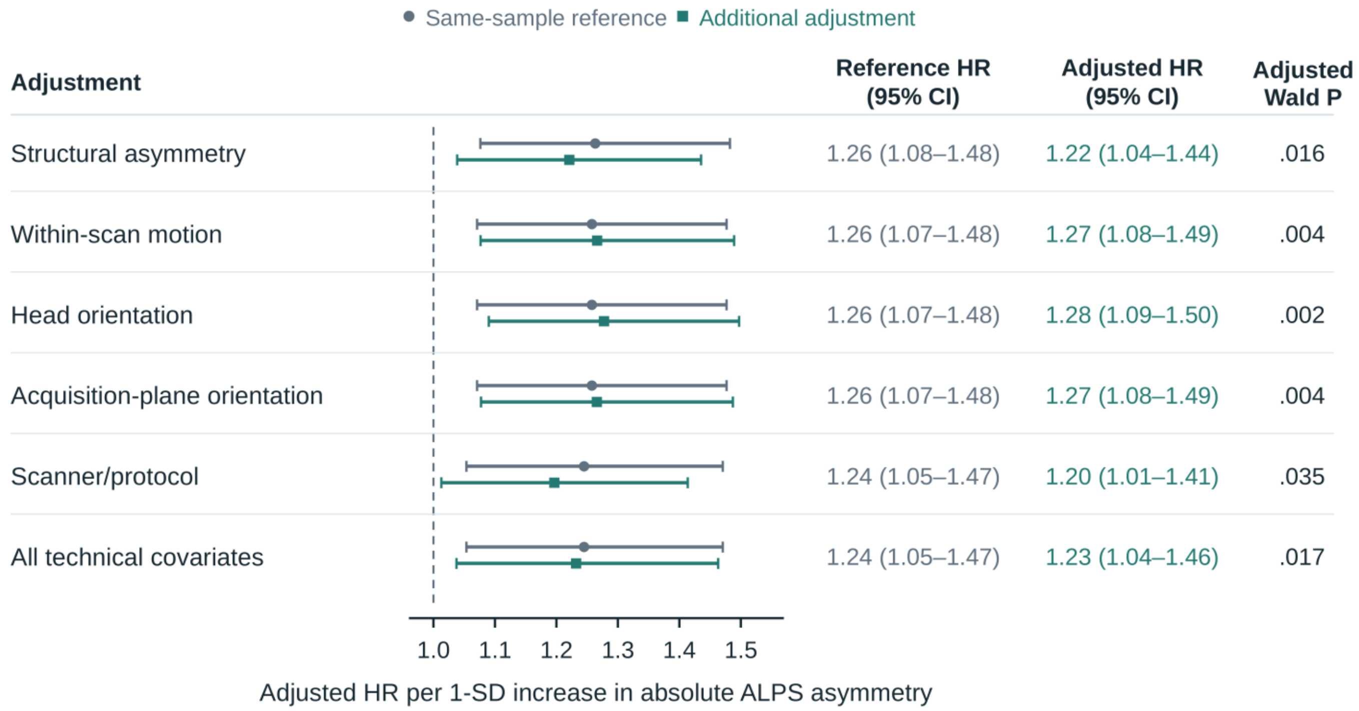


**Figure S1. Sensitivity analyses for the structural asymmetry and technical covariates**. Hazard ratios for AD conversion per 1 SD increase in absolute ALPS asymmetry, with 95% CIs. Gray circles show the reference model refitted in the same complete-case sample as each adjusted model, and teal squares show the estimate after additional covariate adjustment. The structural model includes ventricular volume and ventricular, cortical gray matter, and cortical white matter asymmetries (n = 1,097, 110 events). The within-scan motion, head orientation, and acquisition-plane orientation models include 1,131 participants and 115 events. The scanner/protocol and all technical covariate models include 936 participants and 105 events. Wald P reports the two-sided Wald P value for absolute ALPS asymmetry in each additionally adjusted model. AD = Alzheimer disease; ALPS = analysis along the perivascular space; CI = confidence interval; HR = hazard ratio; SD = standard deviation.
